# Longitudinal Analysis of the Prevalence and Incidence of Obesity-Related Comorbidities and Associated Outcomes: An Electronic Health Record Study in Patients with Overweight and Obesity

**DOI:** 10.64898/2025.11.29.25341251

**Authors:** Zeshui Yu, Yuqing Chen, Hongyi Zou, Manling Zhang, Ning Feng, Yu Chen, Lirong Wang

## Abstract

**Background:** Overweight and obesity are increasing alarmingly in the United States and most substantially increases the burden of obesity-related comorbidities (ORCs). Understanding the prevalence, incidence, and long-term outcomes of ORCs is critical to addressing this public health challenge.

**Methods:** A retrospective cohort study was conducted using electronic health records (EHR). A total of 100,000 patients, consisting of 25,000 from each of the four Body Mass Index (BMI) categories, were randomly selected based on their initial BMI from the University of Pittsburgh Medical Center EHR system. Demographic and clinical characteristics were compared using chi-square tests and ANOVA. The prevalence, incidence, hospitalization rates of ORCs by BMI categories were analyzed. Multivariable Cox regression models were employed to identify risk factors associated with mortality.

**Results:** We identified 87,010 eligible patients. A higher prevalence of heart failure (HF), chronic kidney disease (CKD), nonalcoholic fatty liver disease/nonalcoholic steatohepatitis (NAFLD/NASH), obstructive sleep apnea (OSA), osteoarthritis (OA), and type 2 diabetes (T2DM) was observed among patients with higher BMI categories compared to those classified as overweight. Patients with class III obesity demonstrated significantly higher incidence rates of CKD, HF, hypertension, OSA, OA, and T2DM than other BMI categories. Additionally, a higher cumulative hospitalization rate and an increased risk of mortality were observed among patients with more severe obesity. The Multivariate Cox regression shows several ORCs are positively associated with post-5-year mortality, with CKD (HR: 1.75, 95% CI: 1.63–1.87), HF (HR: 1.68, 95% CI: 1.57–1.80), and T2DM (HR: 1.65, 95% CI: 1.57–1.73) identified as the strongest predictor.

**Conclusion:** Our retrospective cohort study elucidates the epidemiology of ORCs across various BMI categories and identifies key risk factors of post-5-year mortality. These findings emphasize the public health burden of obesity and ORCs and highlight the critical need for integrating comprehensive obesity care into clinical practice according to the severity of obesity.

## Introduction

Obesity is characterized by an excessive accumulation of body fat and has been formally recognized as a disease by the World Health Organization (WHO).[1] However, the prevalence of obesity among adults in the US older than 20 has risen from 30.5% in 1999-2000 to 41.9% in 2017-2020, according to the Centers for Disease Control and Prevention (CDC).[2] The increase in the prevalence of obesity has become one of the most common, significant, and severe health problems in the United States and many other developed countries.[3–5] Body Mass Index, defined as the ratio of body weight (kg) to the square of body height (m^2^), is commonly used to determine the severity of obesity in a clinical setting.[1] Individuals with overweight or obesity were categorized into four groups according to the WHO classification.[1] As described by WHO, overweight is defined as a BMI greater or equal to 25 to 29.9 kg/m^2^, whereas obesity classes I, II, and III are described as having a BMI between 30 and 34.9 kg/m^2^, 35 and 39.9 kg/m^2^, and over 40 kg/m^2^, respectively.[5]

Despite recommendations for weight loss counseling and referrals to lifestyle intervention programs as key components of obesity management guidelines, it has been reported that less than 40% of patients who are overweight or obese receive counseling or education related to weight reduction in primary care settings. [6, 7] Extensive research has established that overweight and obesity are significant contributors to the presence and development of comorbidities, which can subsequently lead to various comorbid conditions and complex downstream health complications. [1, 8–10] Common obesity-related comorbidities (ORCs) include type 2 diabetes mellitus (T2DM), hypertension (HTN), hyperlipidemia (HLP), and coronary artery disease (CAD). The burden of ORCs negatively impacts patients’ quality of life, increases morbidity and mortality, and escalates healthcare expenditures. [10–12] Like chronic conditions such as HTN and T2DM, the management of obesity typically requires lifelong interventions to treat both obesity and its complications, thereby reducing associated health risks.[13]

However, several barriers and challenges hinder the successful integration of weight management strategies into clinical practice, leading to obesity being one of the most commonly underdiagnosed and undertreated diseases. Optimal and personalized management strategies are still impeded by numerous unanswered questions, including targeted approaches to manage patients experiencing different levels of obesity severity. Therefore, it is crucial for researchers to provide descriptive epidemiological electronic health record data collected from real-world clinical settings to assess the burden of obesity-ORCs and associated outcomes across various levels of obesity severity. These insights from real-world evidence can help identify risk patterns and inform more tailored and effective prevention and intervention strategies for patients across different BMI categories.

Our study primarily aims to determine the clinical prevalence and cumulative incidence of ORCs over 5 years in a large random sample of patients managed at the University of Pittsburgh Medical Center (UPMC). Additionally, our secondary objective is to examine the association between BMI categories and comorbidities across various clinical outcomes over time.

## Method

### Data source

The Electronic Health Record (EHR) is considered a promising tool for healthcare professionals to integrate weight loss consultation and formulate adequate management plans to address obesity and ORCs in clinical practice.[14] Neptune is one of the large-scale EHR systems that was used to create a random sample of patients in our study.[15] An EHR is a digital representation of the comprehensive documentation of a patient’s medical history, including numerous types of data, ranging from clinical notes and diagnostic results to medications and treatment plans, capturing detailed and longitudinal patient health information. This data is sourced from various clinical encounters across the healthcare network, allowing for a robust analysis of patient outcomes over time. For this study, de-identified data from the Neptune EHR system was used to ensure patient privacy. The study protocol was approved by the Institutional Review Board (IRB) at the University of Pittsburgh, following all ethical guidelines for human subject research.

### Study cohort

We constructed the study cohort for this retrospective cohort study using patients from the UPMC EHR system, covering the period from January 1st, 2004, to January 31st, 2021. This study examines the medical histories of a randomly selected sample of 100,000 patients aged 18 years or older. Participants with an **initial BMI** on **an entry date** fell into one of four WHO BMI classification groups (27-30 kg/m^2^, 30-35 kg/m^2^, 35-40 kg/m^2^, ≥40 kg/m^2^), with 25,000 patients in each group. Each patient was followed for a minimum of 5 years after their initial measurement (**defined as the observation phase**). This study aims to reflect the epidemiology of obesity and ORCs in real-world clinical settings; therefore, we implemented relatively broad exclusion criteria. To ensure complete documentation of variables, individuals who did not have at least one year of documented medical history prior to the entry date were excluded (n=10,178). Additionally, individuals with less than 5 years of follow-up were excluded due to data entry errors in the EHR (n=2,812).

### Study variables

Initial BMI is defined as the BMI value recorded at the start of the 5-year observation phase. (Fig. 1) Each patient’s age was calculated by subtracting their date of birth from the entry date. Race and ethnicity information was obtained from the demographic files of the EHR. Through a comprehensive literature review, we identified a list of ORCs that includes atrial fibrillation (AFIB), coronary artery disease (CAD), atherosclerotic cardiovascular disease(ASCVD), heart failure (HF), ischemic stroke (IS), myocardial infarction (MI), peripheral artery disease (PAD), chronic kidney disease (CKD), hyperlipidemia (HLP), hypertension (HTN), nonalcoholic fatty liver disease and nonalcoholic steatohepatitis (NAFLD-NASH), obstructive sleep apnea (OSA), osteoarthritis (OA), and type 2 diabetes (T2DM). These comorbidities were identified in the EHR using the International Classification of Diseases-9 (ICD-9) and ICD-10 codes documented at any point up to the end of the 5-year observation phase. A complete list of ICD codes is provided in the supplementary material. To determine what encounter types are qualified as hospitalization, we selected those documented as inpatient or those recorded as hospital encounters associated with a discharge summary to ensure the patient is formally admitted to the hospital to receive inpatient healthcare service.

**Figure 1.**
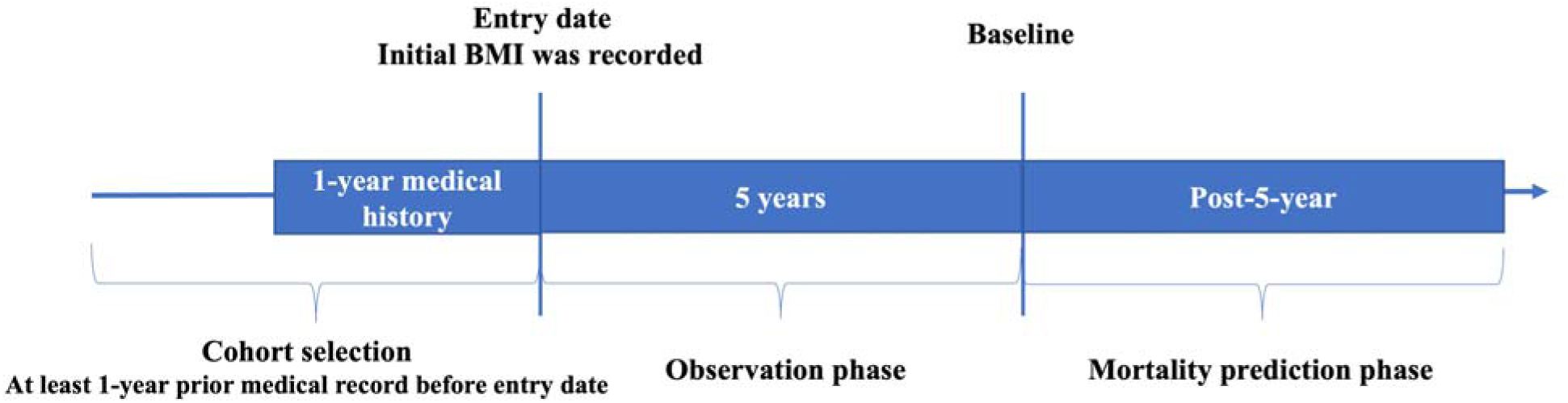
Study Design.

### Statistic methods

Initially, we compared the summary statistics for the demographic and clinical characteristics of our study cohort across BMI categories, using t-tests for continuous variables and ANOVA for categorical variables.

We measured the prevalence and incidence rates of comorbidities by BMI classifications in our study sample. Prevalence of comorbidities at the baseline was defined as the number of patients who had the disease before the entry date, divided by the total number of patients, reflecting the proportion of patients with existing diseases at baseline. In contrast, the cumulative incidence rate was calculated overall and within each specific follow-up period during the observation phase. The incidence rate is calculated by dividing the number of new cases of the disease by the total person-year of patients at risk *1000.

Similarly, the cumulative number of hospitalizations within specific follow-up periods (every half year) was calculated across different BMI categories during the observation phase. This calculation was done by dividing the cumulative number of hospitalizations by the number of patients in each BMI category.

Additionally, we assessed the risk associated with increasing post-5-year mortality among patients who survived the initial 5-year observation phase. (Fig. 1) A multivariable Cox regression model was used to identify risk factors for mortality in our sample of patients with overweight and obesity. The **baseline** was defined as the end of the initial 5-year observation phase, and patients were followed until death, their last encounter, or March 31st, 2021, whichever occurred first (**defined as the mortality prediction phase)**. In our model, the outcome was defined as all-cause mortality, recorded as a single event in the demographic file. The variables included in our model were age, gender, initial BMI categories and all 14 prevalent ORCs at the **baseline**. A two-sided p-value of less than 0.05 was considered statistically significant. All data analyses were conducted using Python 3.9.

Figure 1. Study design flowchart. The flowchart illustrates the process of cohort selection, observation phase, and mortality prediction phases. Patients with an initial BMI recorded on the entry date were categorized into WHO BMI classification groups. Cohort selection required at least one year of prior medical history before the entry date.

## Result

### Patients’ demographics and prevalence of obesity-related comorbidities

A total of 87,010 patients with a minimum of five years of follow-up were included based on the inclusion and exclusion criteria from our random sample for this study. Among these eligible patients, 21,847 had an initial BMI in the overweight category, 21,403 in obesity class I, 22,132 in obesity class II, and 21,628 in obesity class III.

Table 1 presents the overall demographics and characteristics of the study cohort across BMI categories. A total number of 87,010 eligible patients were included in this analysis. Among these individuals, 21,847 (25.11%) patients were classified as having overweight, 21,403(24.60%) as Obesity Class I, 22,132 (25.44%) as Obesity Class II, and 21,628 (24.86%) as Obesity Class III based on their recorded BMI. All associations between patient characteristics and BMI categories were statistically significant (p < 0.05). The mean age of the study cohort was 55.01 years (SD = 15.01). The mean age decreased from 58.42 years in the overweight group to 50.15 years in individuals classified as Obesity Class III. In the entire study cohort, the majority of patients were female (60.81%) and white (86.93%). The proportion of female patients increased from 52.13% in the overweight category to 72.91% in Obesity Class III. Higher BMI categories had a larger proportion of African American individuals, while this pattern was not observed among white patients and those of other races.

**Table 1.**
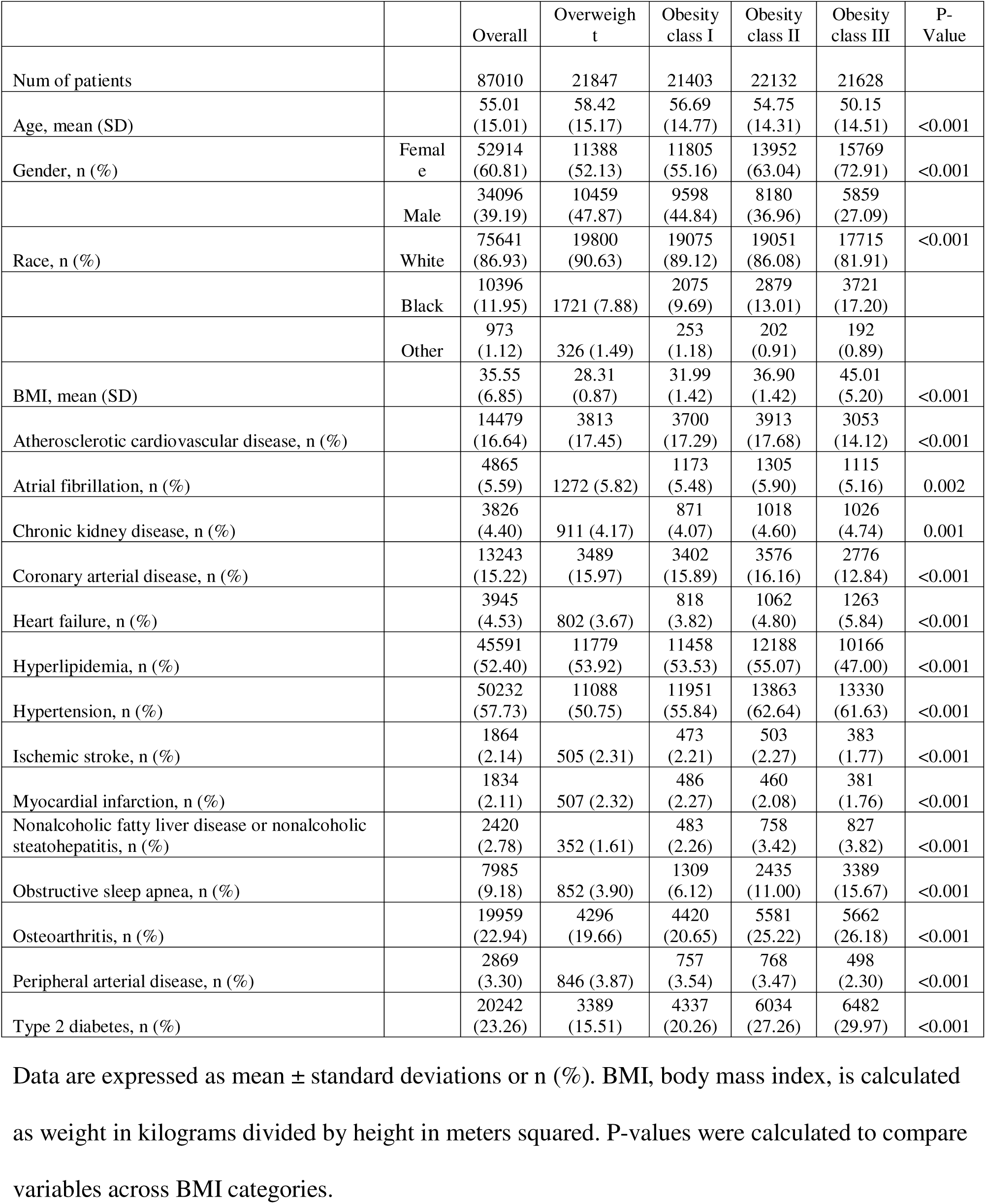
Baseline Demographics and Patient Characteristics of the Study Sample Stratified by BMI Categories.

Table 1 also displays the prevalence of ORCs at the entry date, stratified by BMI categories. Every observed association between the weight categories and disease prevalence was found to be statistically significant. Specifically, the prevalence of HF, NAFLD-NASH, OSA, OA, and T2DM increased as BMI categories rose, with percentages growing from 3.67%, 1.61%, 3.90%, 19.66%, and 15.51%, respectively, in the BMI 27–30 category to 5.84%, 3.82%, 15.67%, 26.18%, and 29.97%, respectively, in Obesity Class III. In Obesity Class II, the highest prevalence was observed for CAD (16.16%), AFIB (5.90%), HLP (55.07%), and HTN (62.64%) compared to other BMI categories. There was a noticeably smaller percentage of patients with ASCVD (14.12%) and HLP (47.00%) in the Obesity Class III category (average percentage of 16.64% and 52.40%, respectively) at baseline. Conversely, the proportion of patients with MI and PAD was with more severe obesity, dropping from 2.32% and 3.87% in the BMI 27–30 category to 1.76% and 2.30% in Obesity Class III.

### The five-year incidence rate of obesity-related comorbidities

This table presents the five-year incidence rates of various obesity-related comorbidities (ORCs) across BMI categories: Overweight (BMI 27-30), Obesity I (BMI 30-35), Obesity II (BMI 35-40), and Obesity III (BMI ≥40). Data are expressed as the number of events/number at risk and incidence rates per 1,000 person-years.

Table 2 displays the five-year incidence rates of various ORCs across different BMI categories during the observation phase. HLP shows the highest incidence rate at 122.19 in patients with obesity II before it decreases to 107.31 in those with obesity III. There is a noticeable increasing trend in the incidence rates of CKD, HF, HTN, NAFLD-NASH, OSA, OA, and T2DM across BMI categories. Specifically, the rates increase from 14.88, 10.79, 79.18, 6.58, 16.45, 58.71, and 15.90 cases per 1,000 person-years in overweight patients to 19.32, 17.69, 129.45, 17.56, 74.00, 81.38, and 49.83 cases, respectively, in Obesity III patients. There are interesting patterns in the incidence rates of ASCVD, CAD, MI, PAD, and AFIB across BMI categories. The incidence rate of ASCVD slightly fluctuates, peaking at 27.78 in Obesity I and then settling at 26.38 in Obesity III. The incidence rate of CAD was observed to increase across BMI categories, reaching a peak of 22.63 in obesity II before slightly decreasing to 22.11 in obesity III. The MI incidence rate was lower in higher obesity categories, with the lowest rate of 5.52 observed in patients with obesity III. Similarly, PAD shows a decreasing trend from a peak of 15.40 in overweight to a low of 12.64 in obesity III. Patients with overweight have been found to have the lowest incidence rate of AFIB; this rate increases slightly with BMI, peaking at 13.17 in obesity II before falling to 13.07 in obesity III.

**Table 2:** Five-Year Incidence Rates of Obesity-Related Comorbidities per 1,000 Person-Years by BMI Category.

|  | Overall |  | Overweight |  | Obesity I |  | Obesity II |  | Obesity III |  |
| --- | --- | --- | --- | --- | --- | --- | --- | --- | --- | --- |
| Obesity-related Comorbidities | No events/No at risk | Incidence rate | No events/No at risk | Incidence rate | No events/No at risk | Incidence rate | No events/No at risk | Incidence rate | No events/No at risk | Incidence rate |
| Atherosclerotic cardiovascular disease | 9110/72531 | 26.86 | 2221/18034 | 26.29 | 2297/17703 | 27.78 | 2298/18219 | 27.01 | 2294/18575 | 26.38 |
| Atrial fibrillation | 5099/82145 | 12.82 | 1245/20575 | 12.49 | 1229/20230 | 12.53 | 1327/20827 | 13.17 | 1298/20513 | 13.07 |
| Chronic kidney disease | 6958/83184 | 17.40 | 1508/20936 | 14.88 | 1678/20532 | 16.99 | 1867/21114 | 18.45 | 1905/20602 | 19.32 |
| Coronary arterial disease | 7679/73767 | 22.04 | 1851/18358 | 21.30 | 1880/18001 | 22.10 | 1979/18556 | 22.63 | 1969/18852 | 22.11 |
| Heart failure | 5435/83065 | 13.52 | 1107/21045 | 10.79 | 1184/20585 | 11.83 | 1419/21070 | 13.92 | 1725/20365 | 17.69 |
| Hyperlipidemia | 17451/41419 | 114.32 | 4122/10068 | 109.81 | 4324/9945 | 119.54 | 4385/9944 | 122.19 | 4620/11462 | 107.31 |
| Hypertension | 14305/36778 | 102.01 | 3454/10759 | 79.18 | 3534/9452 | 96.95 | 3477/8269 | 114.01 | 3840/8298 | 129.45 |
| Ischemic stroke | 3594/85146 | 8.59 | 932/21342 | 8.90 | 969/20930 | 9.44 | 892/21629 | 8.40 | 801/21245 | 7.66 |
| Myocardial infarction | 2578/85176 | 6.13 | 635/21340 | 6.03 | 691/20917 | 6.70 | 672/21672 | 6.29 | 580/21247 | 5.52 |
| Nonalcoholic fatty liver disease or nonalcoholic steatohepatitis | 4958/84590 | 12.06 | 696/21495 | 6.58 | 1014/20920 | 9.92 | 1495/21374 | 14.48 | 1753/20801 | 17.56 |
| Obstructive sleep apnea | 13855/79025 | 38.78 | 1656/20995 | 16.45 | 2569/20094 | 27.39 | 4046/19697 | 46.34 | 5584/18239 | 74.00 |
| Osteoarthritis | 19626/67051 | 69.78 | 4455/17551 | 58.71 | 4685/16983 | 64.83 | 5209/16551 | 76.32 | 5277/15966 | 81.38 |
| Peripheral arterial disease | 5795/84141 | 14.22 | 1562/21001 | 15.40 | 1513/20646 | 15.18 | 1421/21364 | 13.72 | 1299/21130 | 12.64 |
| Type 2 diabetes | 9303/66768 | 30.23 | 1406/1845<br>8 | 15.90 | 1901/1706<br>6 | 23.78 | 2683/1609<br>8 | 36.82 | 3313/1514<br>6 | 49.83 |

### The Cumulative Number of Hospitalizations across BMI Categories

Table 3 presents an analysis of cumulative hospitalization rates over a five-year observation phase across various BMI categories. Hospitalization rates for participants with overweight began at 0.12 hospitalizations per person half a year after the entry date, gradually increasing to

**Table 3.** Cumulative Hospitalizations Over 5 Years Across Different BMI Categories.

| Year | 0.5<br>years | 1<br>year | 1.5<br>years | 2<br>years | 2.5<br>years | 3<br>years | 3.5<br>years | 4<br>years | 4.5<br>years | 5<br>years |
| --- | --- | --- | --- | --- | --- | --- | --- | --- | --- | --- |
| Overweight | 0.12 | 0.21 | 0.30 | 0.39 | 0.50 | 0.60 | 0.70 | 0.82 | 0.93 | 1.06 |
| Obesity class I | 0.13 | 0.22 | 0.32 | 0.43 | 0.53 | 0.64 | 0.75 | 0.87 | 1.00 | 1.13 |
| Obesity class II | 0.15 | 0.26 | 0.38 | 0.5 | 0.63 | 0.75 | 0.88 | 1.02 | 1.17 | 1.32 |
| Obesity class III | 0.17 | 0.3 | 0.44 | 0.58 | 0.73 | 0.89 | 1.05 | 1.22 | 1.39 | 1.57 |

Cumulative hospitalization rates are presented for BMI categories: Overweight (BMI 27–30), Obesity I (BMI 30–35), Obesity II (BMI 35–40), and Obesity III (BMI $\geq 40$ ). Rates were calculated by dividing the cumulative number of hospitalizations by the number of patients in each BMI category at the corresponding follow-up periods.

1.06 hospitalizations per person by the end of the fifth year. For patients classified in Obesity class I category, the initial rate was slightly higher at 0.13, rising to 1.13 after five years. In the Obesity class II group, hospitalization rates started at 0.15 hospitalizations per person and increased more steeply to 1.32 by the fifth year. The most pronounced increase was observed in the Obesity class III group, where the rate started at 0.17 hospitalizations per person and surged to 1.57 hospitalizations per person, indicating a significant correlation between higher BMI and increased hospitalization rates.

### The Predictors for Post-5-year Mortality

This table presents hazard ratios (HRs) with 95% confidence intervals (CIs) for predictors of post-5-year mortality. Predictors include demographic variables (age, gender, race), BMI categories (Overweight [reference group], Obesity Class I, II, III), and obesity-related comorbidities. An HR greater than 1 indicates an increased mortality risk, while an HR less than 1 indicates a decreased risk. P-values reflect the significance of each predictor in the model.

Table 4 displays the results of a multivariate Cox regression analysis evaluating risk factors of post-5-year mortality across BMI categories during the mortality prediction phase. Age and gender were significant risk factors, with each additional year of age increasing mortality risk by 7% (HR: 1.07, 95% CI: 1.07–1.08, p<0.005), and males exhibiting a 32% higher mortality risk than females (HR: 1.32, 95% CI: 1.26–1.38, p<0.005). We identified obesity II and obesity III as significant risk factors compared to overweight. While Obesity I showed no significant effect on mortality (HR: 1.03, 95% CI: 0.97-1.09, p=0.38), BMI Class II and Class III have the HR of 1.13 (95% CI: 1.06 - 1.21, p<0.005) and 1.57 (95% CI: 1.46 - 1.68, p<0.005), respectively. Several ORCs were significantly associated with an increased risk of post-5-year mortality, with CKD (HR: 1.75, 95% CI: 1.63–1.87, p<0.005), HF (HR: 1.68, 95% CI: 1.57–1.80, p<0.005), and T2DM (HR: 1.65, 95% CI: 1.57–1.73, p<0.005) showing the greatest impact. Additional conditions linked to higher mortality risk included CAD, IS, PAD, AFIB, HTN, NAFLD/NASH, and OSA. Interestingly, HLP was found to decrease the risk of mortality, with an HR of 0.76 (95% CI: 0.73-0.81, p<0.005).

**Table 4.**
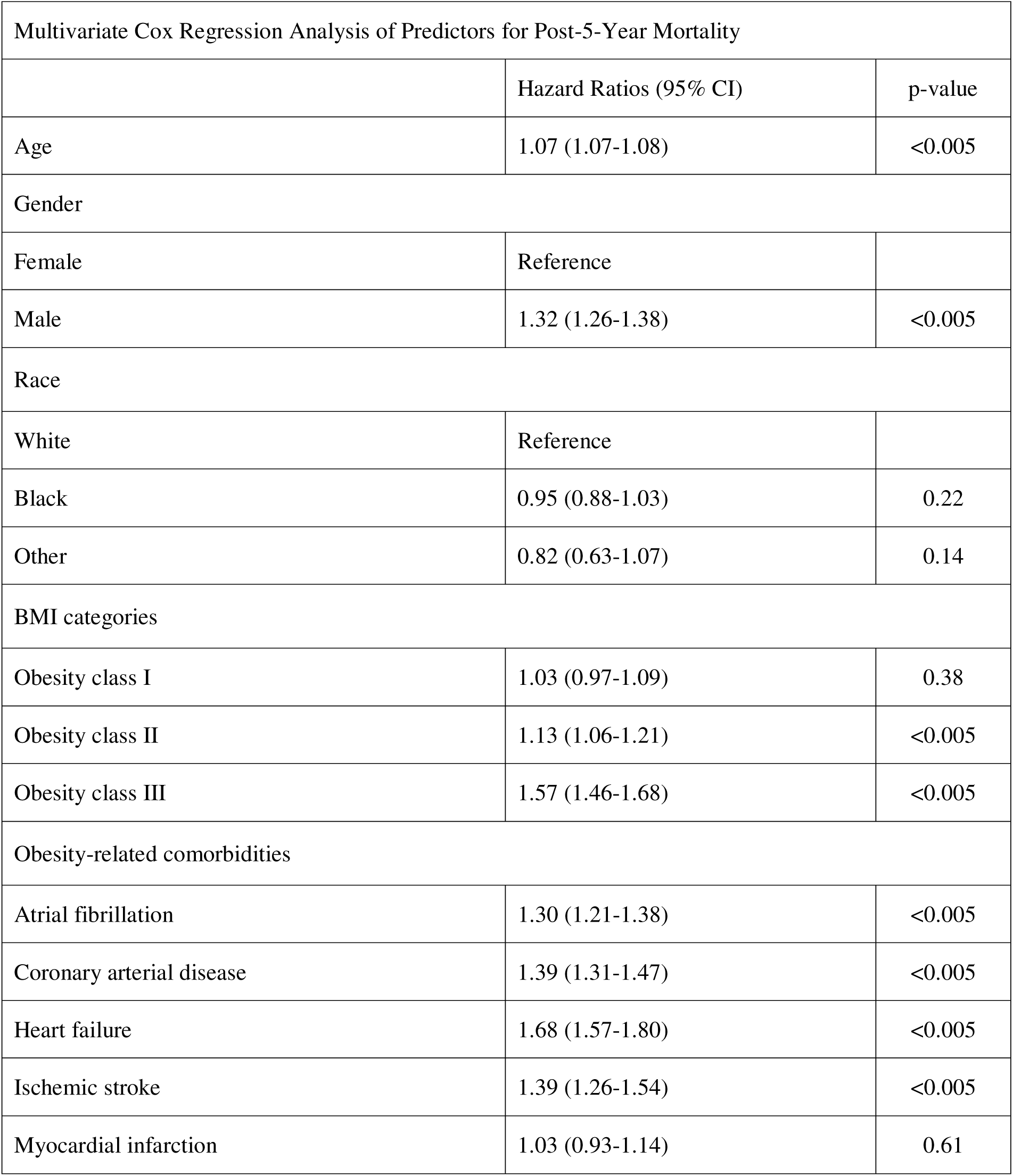
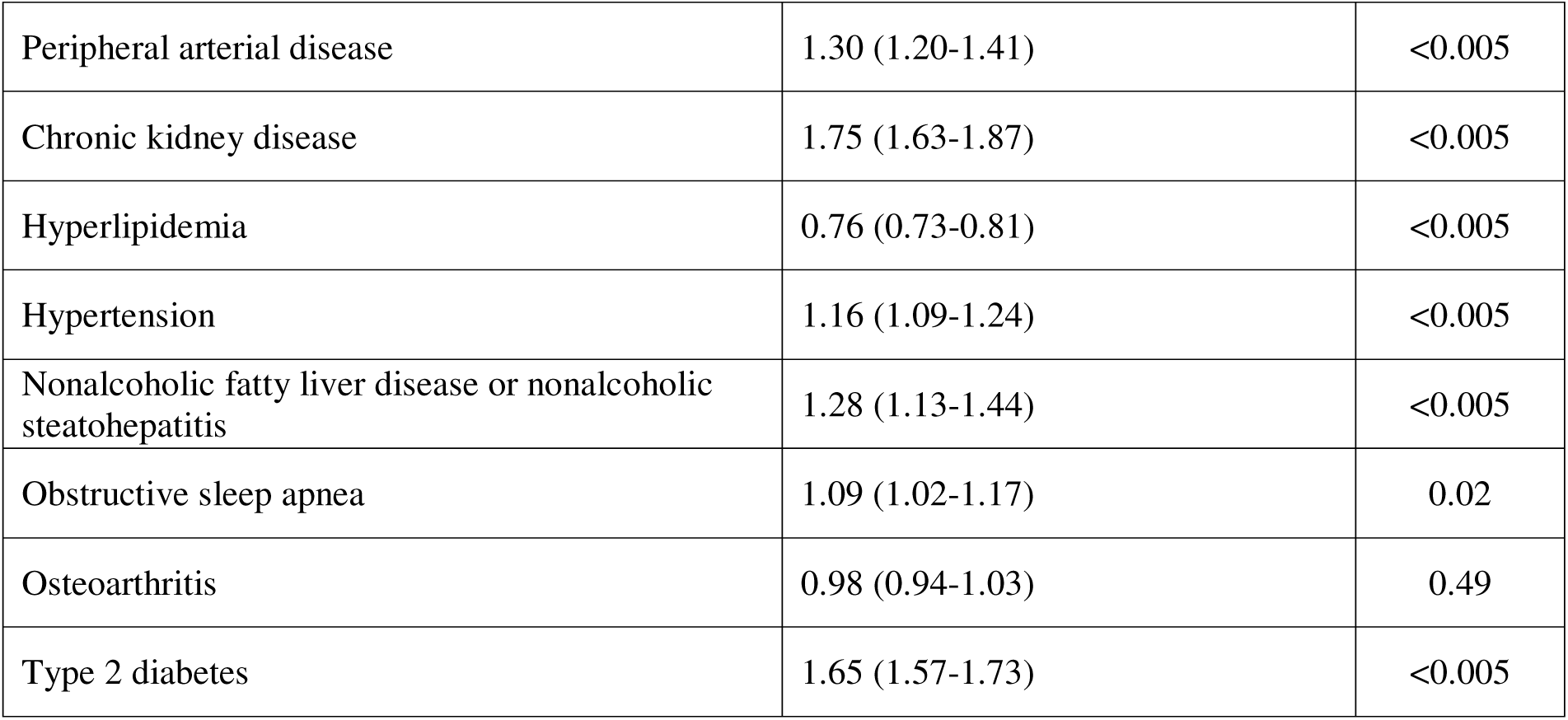
Multivariate Cox Regression Analysis of Predictors for Post-5-Year Mortality.

## Discussion

We conducted a comprehensive retrospective cohort study using a sample of overweight and obese individuals to collect a broad range of information on ORCs across varying obesity severity in clinical settings. First, our findings revealed that the prevalence and incidence of most metabolic ORCs are highest in patients with obesity category III, while the prevalence and incidence of most cardiovascular ORCs are greatest in patients with obesity category II. Second, we observed a positive trend of increasing hospitalizations across higher BMI categories during a 5-year observation phase. Finally, we found that obesity is associated with an increased risk of post-5-year mortality, with age, male gender, obesity severity, and comorbidities such as CKD, HF, and T2DM identified as independent predictors of mortality risk.

Previous population-based studies using the National Health and Nutrition Examination Survey (NHANES) data across the US found that obesity and severe obesity rates increased among adults from 1999 to 2018.[16, 17] Given the epidemic of overweight and obesity, we focused on a large cohort of overweight and obese patients managed within one of the largest healthcare systems in the U.S. Our research provides essential epidemiological data on the burden of ORCs in patients with overweight or obesity across different obesity severity. In our random sample, we observed a higher percentage of women, with a trend showing that they are more likely to have severe obesity compared to men. We also found that obesity class III had a higher proportion of African American patients and a younger average age compared to other categories. This trend largely aligns with the prevalence of obesity and severe obesity reported by the CDC, except for a notable difference in the percentage of women and men, which may be partially explained by women’s more frequent healthcare utilization compared to men. [18, 19]

Overweight and obesity are well-established risk factors for pre-diabetes and T2DM, and screening for T2DM is recommended for all patients with obesity. [20, 21] The rapid increase in obesity prevalence plays a significant role in contributing to the growing number of individuals with type 2 diabetes. [22] A previous study shows that individuals with obesity have a sevenfold higher risk of developing diabetes compared to those of healthy weight and a threefold increased risk compared to overweight individuals. [23] In our cohort, the prevalence of T2DM was 23.26%, and the incidence of T2DM was 30.23 new per 1000 person-year during the 5-year observation phase. A higher prevalence and incidence of T2DM was observed in patients with greater obesity categories. A previous trend analysis among US adults found that the prevalence of total diabetes, defined as having been diagnosed by a doctor or other health professional or having HbA1c levels ≥6.5%, increased from 13.8% in 1999 to 21.3% in 2016 among obese patients.[24] In our overweight and obese cohort, we used the ICD-9 and ICD-10 codes to identify T2DM, and a similar prevalence was present in our cohort.

Both obesity and T2DM are well-described risk factors for various cardiovascular diseases and are strongly associated with other chronic conditions,[25] including CKD,[26] HLP,[27] HTN,[28] NAFLD-NASH,[29] and OSA.[30] In our overweight and obesity cohort, more than half of patients living with HTN and HLP at the initial entry date, with prevalence rates of 52.40% and 57.73%, respectively. The 5-year incidence rates of HTH and HLP were also observed to be the highest among the ORCs analyzed in our study. Our findings correlate with other large-scale EMR studies on healthcare expenditures for ORCs, which indicate that hypertensive diseases and dyslipidemia show the highest annual prevalence among 21 ORCs in obese patients, with 48.30% and 45.10%, respectively. [31] By collecting samples across each BMI category, our study provides epidemiological evidence that the prevalence and incidence of metabolic diseases, such as CKD, NAFLD-NASH, and OSA, are generally higher in patients with severe obesity than those with overweight. The prevalence and incidence of OA, one of the costliest ORCs, was also significantly higher in patients with severe obesity compared to those who were overweight.

Furthermore, we investigated a broad spectrum of cardiovascular diseases, including AFIB, CAD, HF, IS, MI, and PAD. A previous cross-sectional study used EHR data to analyze 324,199 patients and reported that the prevalence of CAD increased from 5.7% in overweight to 6.3% in obesity I patients. The prevalence of HF demonstrated an increased trend across the BMI categories, from 1.5% in overweight to 2.4% in obesity III patients, this increasing trend was also found in our findings. [32] An interesting observation from our study is that, except for HF, there appears to be a slight decrease in the prevalence and incidence rates of other cardiovascular diseases among patients in obesity category III. This may be due to our inclusion criteria, where we required patients to survive the 5-year observation phase. Since cardiovascular diseases often present as acute and potentially fatal events in clinical settings, our cohort may be relatively less sick compared to the general population of overweight or obese patients with cardiovascular diseases. [33, 34] Further research is required to clarify the causal relationship between obesity and cardiovascular disease.

While obesity is recognized as a chronic disease and counseling is recommended in clinical guidelines, unlike ORCs, obesity itself is rarely managed and treated, and the optimal integration of obesity care remains hindered by several challenges.[35, 36] Our study validated a higher rate of hospitalization over a 5-year period among patients with higher BMI levels, as previously reported in a large academic setting, with 13 years of follow-up, the covariate-adjusted average number of all-cause hospitalization increase across the normal weight, overweight, and obese adults.[37] Frequent hospitalizations of patients with obesity provide significant opportunities for clinicians to engage these individuals, effectively implement obesity management plans, control modifiable risk factors, enhance health outcomes, and reduce the public health cost and burden of obesity. The successful integration of obesity management and initiation of appropriate therapies requires enhanced healthcare provider training,[38] increased awareness,[39] and reduced stigma,[40] collectively paving the way for more effective and proactive healthcare interventions.

Leveraging the longitudinal data from our cohort, our study is the first to examine the association between potential risk factors and post-5-year mortality in patients with obesity. Patients in obesity Classes II and III exhibited a 13% and 57% higher risk of mortality after five years, respectively, compared to those in the overweight category. These findings corroborate and extend existing evidence that more severe obesity significantly elevates mortality rates, demonstrating that the temporal effects of obesity persist beyond a five-year timeframe.[41, 42] Additionally, we have identified a list of risk factors such as AFIB, CAD, HF, IS, MI, PAD, CKD, HLP, HTN, NAFLD-NASH, OSA, OA, and T2DM for post-5-year mortality in overweight and obese populations. HLP is associated with a reduced risk of mortality post-five years, likely due to effective management of the condition and the mortality-lowering benefits of statin use in this population.[43, 44]

The primary strength is that our study systematically characterizes the health status with comprehensive longitudinal follow-up based on EHR data, which differs from community-dwelling datasets. Providing the most recent trends and statistics on obesity within the medical system, our findings offer a unique and detailed perspective on longitudinal epidemiology data and ORCs stratified by categorical BMI class. This also allowed us to provide examinations of how the impact of the severity of obesity on health outcomes. Furthermore, unlike studies based on surveys or self-reported metrics, our research benefits from weight and height measurements taken by trained clinicians, ensuring accurate BMI calculations. This, along with diagnoses of ORCs by healthcare professionals, provides enhanced accuracy and richer clinical insights into obesity management.

Certain limitations of the current study should be noted. First, it is a descriptive observational study, establishing causality is challenging. Further research is needed to explore the causal relationship between obesity status and its comorbid medical conditions and health outcomes. Second, we included a unique sample of overweight and obese subjects. Our cohort experiences a higher burden of comorbidities compared to the general population of overweight and obese individuals who do not require constant management or seek health care in our healthcare institution. However, they are healthier than the typical hospital-managed patients when considering a minimum five-year survival without any severe or acute illness leading to mortality. Therefore, generalizing our findings may present challenges. Third, data entry errors are common in EHR due to billing priorities and time constraints, which could lead to delayed or underdiagnosed conditions. Given the large sample size of our study, the potential impact of inaccurate coding is likely to be diminished.

## Conclusion

The results of this retrospective cohort study reveal the health status of patients with overweight or obesity across various BMI categories, showing a higher prevalence and incidence of ORCs and elevated hospitalization rates in patients with obesity compared to those who are overweight. Additionally, we observed the long-term effects of the severity of obesity and a range of risk factors on post-five-year mortality. Our study highlights the escalating public burden of obesity epidemiology and underscores the necessity for the successful integration of obesity care in clinical settings.

## Data Availability

All data produced in the present study are available upon reasonable request to the authors

## List of Abbreviations

AFIB: Atrial Fibrillation
ANOVA: Analysis of Variance
ASCVD: Atherosclerotic Cardiovascular Disease
BMI: Body Mass Index
CAD: Coronary Artery Disease
CKD: Chronic Kidney Disease
EHR: Electronic Health Records
HF: Heart Failure
HLP: Hyperlipidemia
HTN: Hypertension
ICD: International Classification of Diseases
IRB: Institutional Review Board
IS: Ischemic Stroke
MI: Myocardial Infarction
NAFLD/NASH: Nonalcoholic Fatty Liver Disease/Nonalcoholic Steatohepatitis
OA: Osteoarthritis
ORCs: Obesity-Related Comorbidities
OSA: Obstructive Sleep Apnea
PAD: Peripheral Arterial Disease
T2DM: Type 2 Diabetes Mellitus
UPMC: University of Pittsburgh Medical Center
WHO: World Health Organization

## Declarations

### Ethics approval and consent to participate

This study was approved by the Institutional Review Board (IRB) at the University of Pittsburgh, and informed patient consent was waived. The study utilized de-identified EHR data to ensure patient privacy.

### Consent for publication

Not applicable.

### Availability of data and materials

The data used in this study were from UPMC under a data use agreement. The authors are not permitted to distribute the data to any third party, but researchers may contact UPMC for data access.

### Authors’ contributions

ZY played a leading role in defining the research question, designing the study, analyzing the data, applying statistical methods, and composing the manuscript while also providing insights throughout the research process. YQ was involved in the study design, statistical method, and manuscript reviewing. HZ contributed to manuscript editing and formatting. LW and YC were involved in the concept, data acquisition, study design, interpretation, comprehensive review and revision of the final manuscript, the final approval of the manuscripts, and project management. MZ and FN engaged in concept, study design, and discussion. FS contributed to the study design, interpretation, discussion, review, and revision of the manuscript and final approval. All authors read and approved the final manuscript.

